# Prevalence of anti-platelet factor4/polyanionic antibodies after COVID-19 vaccination with ChAdOx1 nCoV-19 and CoronaVac in Thais

**DOI:** 10.1101/2021.07.15.21260622

**Authors:** Phichchapha Noikongdee, Pornnapa Police, Tichayapa Phojanasenee, Pichika Chantrathammachart, Pimjai Niparuck, Teeraya Puavilai, Angsana Phuphuakrat, Pantep Angchaisuksiri, Kochawan Boonyawat

## Abstract

**Introduction:** Vaccine-induced thrombotic thrombocytopenia (VITT) has been reported after vaccination with the adenoviral vector COVID-19 vaccine ChAdOx1 nCoV-19 in European countries. To date, no case of VITT has been reported in Thais after COVID-19 vaccination. We determined the frequency of anti-PF4/polyanionic antibodies in the Thai population receiving the COVID-19 vaccines.

**Methods:** We conducted a cross-sectional study to evaluate the prevalence of anti-PF4/polyanionic antibodies in health care workers who received COVID-19 vaccination with ChAdOx1 nCoV-19 or CoronaVac within 7-35 days. A control population who had not been vaccinated was also included. Anti-PF4/polyanionic antibodies were detected using enzyme-link immunosorbent assay (ELISA). Functional assay with platelet aggregation was performed for all positive anti-PF4/polyanionic antibody ELISA tests.

**Results:** A total of 646 participants were included in the study. 221 received ChAdOx1 nCoV-19, 232 received CoronaVac, and 193 participants were in the control group. The prevalence of anti-PF4 antibodies was 2.3% (95% confidence interval [CI] 0.7 to 5.2), 1.7% (95% CI, 0.5 to 4.4) in the ChAdOx1 nCoV-19 and CoronaVac groups, respectively. There was no positive test in the control group. None of the PF4/polyanionic positive sera induced platelet aggregation.

**Conclusion:** We found a low prevalence of anti-PF4 antibodies in Thais after vaccination with ChAdOx1 nCoV-19 and CoronaVac. Low titer positive PF4/polyanionic ELISA results should be interpreted with caution when screening asymptomatic individuals after vaccination against COVID-19.

**Highlights:**

- High titer PF4/polyanion antibodies are associated with vaccine-induced immune thrombotic thrombocytopenia (VITT).
- Low titer PF4/polyanion antibodies occur after vaccination with ChAdOx1 nCoV-19 and CoronaVac in some vaccinees.
- These PF4/polyanion antibodies do not activate platelets and lack of association with VITT.
- Low-titer positive PF4/polyanionic ELISA results should be interpreted with caution when screening asymptomatic individuals after vaccination against COVID-19.

## Introduction

Coronavirus disease 2019 (COVID-19) has resulted in devastating worldwide morbidity and mortality since late 2019. Vaccination against COVID-19 is a cornerstone measure in the control of the COVID-19 pandemic. Currently, ChAdOx1 nCoV-19 (AstraZeneca) and CoronaVac (Sinovac Biotech) are the only 2 available vaccines in Thailand. Recent reports of vaccine-induced thrombotic thrombocytopenia (VITT) after ChAdox1 nCoV-19 vaccination have raised concern of using the COVID-19 vaccine [1-3]. Almost all of the cases presented with thrombosis in unusual sites, thrombocytopenia and had a strongly positive anti-platelet factor4 (PF4)/polyanionic antibodies. The syndrome resembles that of autoimmune heparin-induced thrombocytopenia (HIT). Several European countries have paused or restricted the use of the ChAdOx1 nCoV-19 vaccine due to the perceived risk of this severe adverse event. To date, no case of VITT has been reported in Thailand after vaccination with ChAdOx1 nCoV-19 of more than 1.7 million doses. Case reports of this syndrome are also sparse in the Asian population. To support the vaccination campaign and to reassure the public about mass vaccination in Thailand, we conducted a cross-sectional study to evaluate the prevalence of post-vaccination anti-PF4/polyanionic antibodies among Thais receiving ChAdOx1 nCoV-19 and CoronaVac in order to identify individuals who may be at risk of developing VITT, and to investigate the prevalence of subclinical anti-PF4/polyanionic antibodies. Enzyme-linked immunosorbent assay (ELISA) has been shown to reliably detect anti-PF4/polyanionic antibodies associated with VITT, and was thus used for antibody screening in this study.

## Materials and Methods

### Study population and settings

A cross-sectional study was conducted at Ramathibodi Hospital, a tertiary care academic hospital in Bangkok, Thailand. Adult health care workers who had received COVID-19 vaccination with either ChAdOx1 nCoV-19 or CoronaVac within 7-35 days were included in the study. Healthy volunteer participants who had not been vaccinated were also included as a control group. All participants gave written informed consent. Baseline characteristics including age and sex were recorded. The study protocol was approved by the Human Research Ethics Committee of the Faculty of Medicine at Ramathibodi Hospital, Mahidol University.

### Blood collection and laboratory analysis

After informed consent, blood was collected in a citrate anticoagulant tube from participants. Anti-PF4/polyanionic antibodies were screened by IgG-specific ELISA (Hyphen Biomed Zymutest HIA IgG, Quadratech Diagnostics, UK) according to the manufacturer’s instructions. Results were interpreted as positive if the optical density (OD) was above 0.3. Positive samples in ELISA were tested by platelet aggregation on the CHRONO-LOG® platelet aggregometer (Chrono-log Corporation, PA, USA). Normal blood group O donor platelets were incubated with PF4/polyanion-positive sera in the presence of low-dose heparin (unfractionated heparin 1.0 IU/mL), high-dose heparin (unfractionated heparin 100 IU/mL), or saline buffer. A previously confirmed HIT serum was used as a positive control, and normal pooled plasma as negative control.

### Statistical analysis

Baseline characteristics were analyzed and presented with mean and standard deviation or median and interquartile range as appropriate. All statical analyses were performed on GraphPad Prism 9.1.1 (GraphPad Software, CA, USA) and Stata statistical software version 15.1 (StataCorp, TX, 2018).

## Results

A total of 453 healthcare workers and 193 controls were included. 221 participants received one dose of ChAdOx1 nCoV-19 and 232 received either one or two doses CoronaVac with an interval of 21±7 days between doses. All participants who received ChAdOx1 nCoV-19 had the first dose of the vaccine. Of the 232 participants receiving CoronaVac, 149 (64.2%) and 83 (35.8%) had the first and the second dose of the vaccine. Median age (interquartile range; IQR) was 61 (38-68), 35 (30-42), 49 (38-55) years in the ChAdOx1 nCoV-19, CoronaVac, and control groups, respectively. Females accounted for 65.6%, 83.6%, and 29.5% in the ChAdOx1 nCoV-19, CoronaVac, and control groups, respectively. The median day (range) post-first vaccination was 23 (18-27) and 18.5 (10-34), respectively (Table 1). No participant had a history of heparin exposure within 3 months.

**Table 1.** Baseline characteristics of study population.

| Characteristics | Vaccine type, *n* (%) |  |  |  |  |
| --- | --- | --- | --- | --- | --- |
|  | Normal control<br>(*n*=193) | ChAdOx1<br>nCoV-19<br>1<sup>st</sup> dose<br>(*n* = 221) | CoronaVac |  |  |
|  |  |  | 1<sup>st</sup> dose<br>(*n* = 149) | 2<sup>nd</sup> dose<br>(*n* = 83) | 1<sup>st</sup>+2<sup>nd</sup> dose<br>(*n* = 232) |
| Median age, years (IQR) | 49<br>(38.0-55.5) | 61<br>(38.0-68.0) | 35<br>(30.0-41.5) | 35<br>(30.0-42.0) | 35<br>(30.0-42.0) |
| Female, *n* (%) | 57 (29.5) | 145 (65.6) | 124 (83.2) | 70 (84.3) | 194 (83.6) |
| Postvaccination<br>PF4/heparin ELISA +, *n*<br>(%) | N/A | 5 (2.3) | 1 (0.7) | 3 (3.6) | 4 (1.7) |
| Median day since first<br>vaccination, days (IQR) | N/A | 23<br>(18.0-27.0) | 12<br>(9.0-17.5) | 36*<br>(33.0-39.0) | 18.5<br>(10.0-34.0) |

\* Median day since second dose vaccination (IQR) was 10 (7.0-17.0) days. N/A; not applicable

Positive anti-PF4/polyanionic antibodies were detected in 5, 4, and 0 samples in the ChAdOx1 nCoV-19, CoronaVac, and control groups, respectively. Therefore, the prevalence of anti-PF4 antibodies was 2.3% (95% confidence interval [CI] 0.7 to 5.2), 1.7% (95% CI, 0.5 to 4.4), and 0% in the ChAdOx1 nCoV-19, CoronaVac, and control groups, respectively. Median OD (range) was 0.03 (0.00-0.70), 0.04 (0.01-0.93), and 0.04 (0.01-0.29) in the ChAdOx1 nCoV-19, CoronaVac, and control groups, respectively. Overall, the results were positive in 9 samples. The mean OD of the positive results was 0.72 (SD 0.17). None of the positive samples showed an OD greater than 1.0. One positive test in the CoronaVac group occurred after receiving the first vaccine dose, and three positive tests after the second dose (Figure 1). None of the PF4/polyanion-positive sera induced platelet aggregation in the Chrono-Log platelet aggregation assay. None of the study participants developed thrombosis or clinically evident VITT.

**Figure 1.**
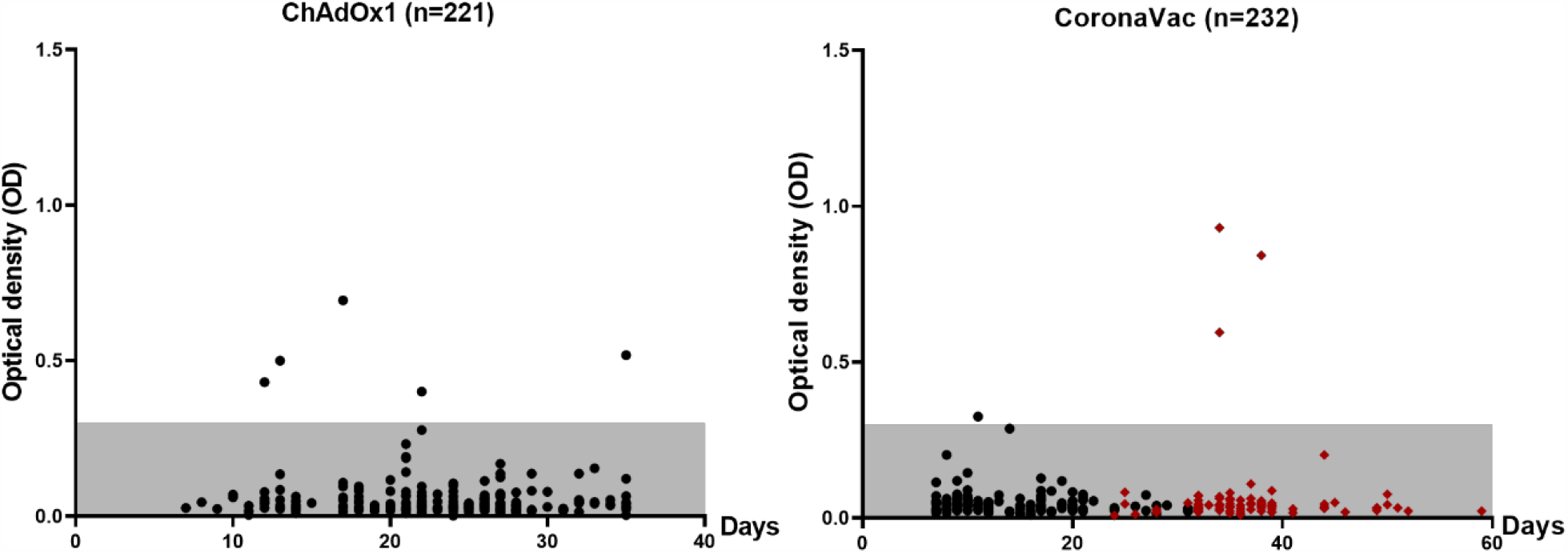
Anti-PF4/polyanionic antibodies were positive in 5 participants in the ChAdOx1 nCoV-19, and 4 participants in the CoronaVac group by ELISA. The highest OD was 0.9. OD of > 0.3 indicates positive result. Black dots indicate OD after first dose vaccination and red diamonds indicate OD after second dose vaccination. OD; optical density. Gray shading indicates negative range.

## Discussion

We report a low prevalence of anti-PF4/polyanionic antibodies in the Thai population receiving COVID-19 vaccination with ChAdOx1 nCoV-19 or CoronaVac. Participants who received ChAdOx1 nCoV-19 and CoronaVac had a similar prevalence of anti-PF4 antibodies, while there was no positive result in the control group. Our data suggest that vaccination against COVID-19 leads to low titer anti-PF4 antibodies in some subjects and that this may occur after vaccination with either ChAdOx1 nCoV-19 or CoronaVac as part of the immunological response.

The results of our study were comparable to a prior study from Norway [4] that reported a 1.2% prevalence of anti-PF4/polyanion antibodies after COVID-19 vaccination with ChAdOx1 nCoV-19. The anti-PF4/polyanion antibodies were also negative in all participants in the control group in that study. However, a study from Germany demonstrated a higher frequency of anti-PF4 antibodies among vaccinees receiving ChAdOx1 nCoV-19 (8.0%) [5]. It also demonstrated seroconversion of anti-PF4 in 2 out of 6 available samples while preexisting antibodies were found in the other 4 available samples. These two studies used different PF4/polyanion ELISA assays. However, both studies also showed that ODs were mostly low (<1.0) and none of the PF4/polyanion ELISA-positive samples induced platelet activation.

There has been no report of VITT after vaccination with CoronaVac, which is an inactivated virus vaccine. However, the prevalences of anti-PF4/polyanionic antibodies after vaccination with ChAdOx1 nCoV-19 and CoronaVac were similar in this study. Low titer positive PF4/polyanionic ELISA results should be interpreted with caution when screening asymptomatic individuals after vaccination against COVID-19. The link between vaccination and antibodies formation is unknown. Sera from patients with clinically-overt VITT are strongly-positive by anti-PF4/polyanion ELISA (typically ODs >2), and cause strong platelet activation in the platelet activation assay.

To date, no case of VITT has been reported in Thailand after vaccination with ChAdOx1 nCoV-19 after more than 1.7 million doses. Case reports of this syndrome are also sparse in the Asian population. It is possible that the incidence of VITT might be lower in Asians. We previously reported a lower prevalence of positive anti-PF4/heparin antibodies and clinical heparin-induced thrombocytopenia after cardiac surgery in Thai patients than in Caucasian patients [6]. Recently it was shown that HLA-DRB1*03:01 and DQB1*02:01 were significant risk factors for increased anti-PF4/heparin antibodies following heparin exposure among inpatients [7]. Frequency distribution of the DRB1*03:01-DQB1*02:01 haplotype across the world varies among different countries (0.1% in Thailand, 12.7% in the UK, and 14.4% in the USA) [8]. Whether this can explain the low prevalence of anti-PF4/heparin antibodies in Thais needs to be further evaluated.

In conclusion, we found a low prevalence of anti-PF4/polyanion antibodies among Thais vaccinated with ChAdOx1 nCoV-19 or CoronaVac. None of the positive antibodies are functional. Low-titer positive PF4/polyanionic ELISA results should be interpreted with caution when screening asymptomatic individuals after vaccination against COVID-19.

## Data Availability

The datasets generated during and/or analysed during the current study are available from the corresponding author on reasonable request.

## Acknowledgements

We thank Assistant Professor Kanlayanee Khupulsup, Department of Pathology, Faculty of Medicine, Ramathibodi Hospital, Mahidol University and all involved personnels who supported in collecting the specimen. We sincerely thank Professor Nigel Key (University of North Carolina) for reviewing the manuscript and offering comments. This study was supported by a medical research grant from Ramathibodi Foundation, the Faculty of Medicine, Ramathibodi Hospital, Mahidol University, Bangkok, Thailand.

## Author’s contribution

P.Noikongdee, P.P., T.Phojanasenee recruited participants, performed the laboratory tests and ran the project. P.P. and K.B. performed the statistical analyses. P.C., T.Puavilai, P.Niparuck, and A.P. collected the specimen. A.P. critically revised the manuscript. P.A. and K.B. designed the study and wrote the manuscript.

